# A Neuro-Symbolic Knowledge Graph and Large Language Model Hybrid Architecture for Multi-Modality Mental Health Counseling

**DOI:** 10.64898/2026.07.29.26359268

**Authors:** Jun Tao, Natalie Fenn, Hannah Parent, Hao Wu, Trisha Arnold, Jennifer Etue, Elizabeth S. Chen, Philip A. Chan

## Abstract

**Background:** Depression and anxiety are managed largely between clinical visits, yet outpatient care lacks scalable, accountable mechanisms for between-visit support. Large language models converse fluently but fuse clinical reasoning with language generation in one opaque process, so they cannot reliably deliver evidence-based psychotherapy and typically operate outside clinician oversight.

**Objective:** To evaluate C-Mind, a provider-supervised neuro-symbolic system in which a Clinical Knowledge Graph (KG) governs therapeutic decisions for a large language model across eight psychotherapy modalities.

**Methods:** Two simulation regimes addressed eight pre-specified governance questions: a *structural validation* of KG routing against 117 guideline-anchored vignettes, and a *governance battery* using progressively disclosing LLM patient agents to evaluate decision traceability, repeatability, provenance auditability, adversarial crisis-detection robustness (277 probes), provider treatment-goal governance, and counselor technique adherence. Crisis detection was additionally validated externally against an independent, clinician-annotated corpus (CRADLE Bench).

**Results:** The KG routed 116/117 vignettes (99.1%) to guideline-appropriate care and detected all 18 high-risk presentations, firing a therapy-suppressing hard halt on 16/18. Adversarial crisis-detection sensitivity was 96.7% and specificity 95.4% (277 probes); on external validation, the system detected 98.5% of 600 dialogues with ongoing suicidal ideation or self-harm at or before the annotator’s confirming turn. Decisions were 99.1% repeatable, 100% reconstructable per turn, and 100% provenance-auditable across all 354 KG nodes. Provider-set diagnosis, goals, and safety context governed behavior deterministically. Stripped of governance, the same model produced unsolicited clinical monologues on 100% of turns (vs 9% governed) and delivered diagnoses and medication advice the governed system never produced.

**Conclusions:** A neuro-symbolic architecture achieves near-perfect guideline-appropriate routing with a governance profile — traceability, reproducibility, machine-traceable provenance, externally validated crisis detection, and deterministic provider control — aligned with requirements for regulated clinical AI.

**Highlights:**

- Clinical Knowledge Graph governs LLM psychotherapy without fine-tuning
- 100% crisis-detection sensitivity across 18 high-risk vignettes
- Per-turn KG decisions fully traceable and reconstructable for audit
- 100% guideline provenance on all 354 nodes; 99.1% five-run repeatability

## 1. Introduction

Depression and anxiety disorders are among the most prevalent and disabling mental health conditions in the United States, affecting approximately 21 million adults with major depressive disorder and 40 million adults with anxiety disorders.[1,2] Although effective psychotherapies, pharmacotherapies, and collaborative care models are available, many individuals do not receive timely, adequate, or continuous treatment.[2] This persistent treatment gap reflects structural constraints in the mental health care system, including clinician workforce shortages, geographic maldistribution of services, prolonged outpatient wait times, and fragmented care delivery.[3]

Even after treatment begins, care remains organized largely around discrete clinical encounters. Psychotherapy visits commonly occur weekly, biweekly, or monthly, leaving extended intervals during which patients are expected to manage symptoms, complete therapeutic assignments, follow behavioral plans, and recognize escalating risk with limited direct clinical guidance.[4] During these intervals, symptom worsening, functional decline, treatment nonadherence, dropout, and safety concerns may emerge before clinicians can intervene.[4–6] Yet routine outpatient care lacks scalable mechanisms for structured monitoring, therapeutic reinforcement, or timely clinician notification between visits. This gap is especially consequential for depression and anxiety, which often require longitudinal symptom monitoring, iterative treatment adjustment, and sustained behavioral reinforcement to achieve and maintain remission.[7]

Digital mental health tools and conversational agents have been proposed as a means of extending support beyond scheduled encounters. Existing approaches include direct-to-consumer chatbots, single-condition digital therapeutics, provider-network platforms, and newer clinician-supervised artificial intelligence (AI) services.[8–15] Direct-to-consumer chatbots generally operate outside clinician workflows and provide limited provider visibility into patient disclosures, symptom trajectories, or emerging safety concerns.[10] Single-condition digital therapeutics typically deliver structured content for a specific disorder or modality, which may limit their applicability to comorbid depression and anxiety presentations.[11–13] Provider-network platforms offer between-session features such as messaging, mood tracking, journaling, and skills practice; however, therapeutic technique selection, escalation, and documentation often remain clinician-mediated rather than governed by auditable AI decision logic.[14,15] Newer clinician-supervised AI services may extend support between visits, but many rely on probabilistic classifiers layered on general-purpose large language models (LLMs) and deliver care through their own clinician networks rather than augmenting the patient’s existing provider.

These limitations are compounded by a central technical challenge in LLM-based mental health tools: conversational output and clinical reasoning are often generated through the same opaque model process.[16,17] Although LLMs can produce fluent and empathic responses, they are prone to hallucination, variability across repeated runs, and limited interpretability, and they do not inherently expose the reasoning used to assess symptom severity, select therapeutic strategies, identify contraindications, or determine whether crisis escalation is needed.[16–18] This opacity limits clinicians’ ability to determine whether a response is consistent with a patient’s treatment plan, why a particular intervention was selected, or whether the same presentation would produce the same decision across independent runs. Recent reports of harms associated with unsupervised AI interactions among emotionally vulnerable users have intensified attention to these risks,[19] and emerging regulatory frameworks increasingly emphasize transparency, clinician reviewability, documentation, risk management, and human oversight for health-related AI systems.[20,21]

Taken together, the between-visit care gap, limitations of existing tools, and governance challenges of LLM-based systems point to the need for a provider-supervised, auditable, multimodal platform for depression and anxiety care. In such a model, the platform does not function as an independent therapist or direct-to-consumer substitute, but as an extension of the clinician’s treatment plan. The supervising clinician defines the patient’s goals, authorized therapeutic approaches, monitoring priorities, and safety parameters. Within these boundaries, the platform can deliver structured between-visit support, reinforce assigned treatment goals, monitor symptoms and engagement, and alert the clinician when predefined safety or deterioration thresholds are met. This model preserves clinician accountability while extending structured support between scheduled encounters.

Reimbursement policy has also begun to recognize between-visit monitoring and care management as reimbursable clinical work when delivered under appropriate supervision and documentation. Behavioral Health Integration, Collaborative Care Management, and Remote Therapeutic Monitoring payment codes support activities such as symptom monitoring, care coordination, digital therapeutic engagement, and clinician review between encounters.[22,23] These frameworks align with core requirements for provider-supervised AI, including treatment-plan alignment, structured monitoring, clinician review, and auditable documentation, thereby providing an implementation pathway for platforms designed to support depression and anxiety care between visits.

To address these needs, we developed C-Mind, a provider-supervised software-as-a-service (SaaS) platform for between-visit depression and anxiety care. C-Mind uses a neuro-symbolic architecture that combines a Clinical Knowledge Graph (KG) with an LLM, separating clinical reasoning from natural language generation.[24,25] The KG encodes eight evidence-based therapeutic modalities: Motivational Interviewing (MI), Cognitive Behavioral Therapy (CBT), Dialectical Behavior Therapy skills (DBT), Acceptance and Commitment Therapy (ACT), Behavioral Activation (BA), Relaxation Techniques (RT), Exposure Therapy (EXP), and Exposure and Response Prevention (ERP). The KG governs therapeutic routing, safety escalation, and technique selection, while the LLM generates patient-facing responses within KG-authorized clinical boundaries. C-Mind is designed to operate within clinician-supervised workflows rather than as a standalone consumer chatbot.

In this study, we conducted a simulation-based evaluation of C-Mind across a pre-specified library of standardized scenarios spanning depression, anxiety, co-occurring presentations, and high-risk cases. We evaluated clinical-domain coverage, guideline-concordant routing, safety escalation, decision repeatability, per-turn traceability, guideline-provenance auditability, treatment-goal alignment, and KG-to-LLM technique enforcement. Together, these analyses assess whether a provider-supervised neuro-symbolic SaaS platform can support accountable between-visit mental health care while preserving transparent and auditable clinical decision logic.

## 2. Methods

### 2.1 System Overview

C-Mind is a HIPAA-compliant, provider-supervised SaaS platform for between-visit depression and anxiety care, deployed on Azure cloud infrastructure. The platform separates clinical reasoning from language generation through three components (Figure 1). The Symbolic Layer processes each patient message through the Clinical KG and produces an explicit clinical directive. The Neural Layer uses an LLM to generate a naturalistic therapeutic response within KG-authorized boundaries. The Post-Response Safety Gate applies deterministic crisis-resource injection and protected health information (PHI) scrubbing before delivery to the patient.

**Figure 1.**
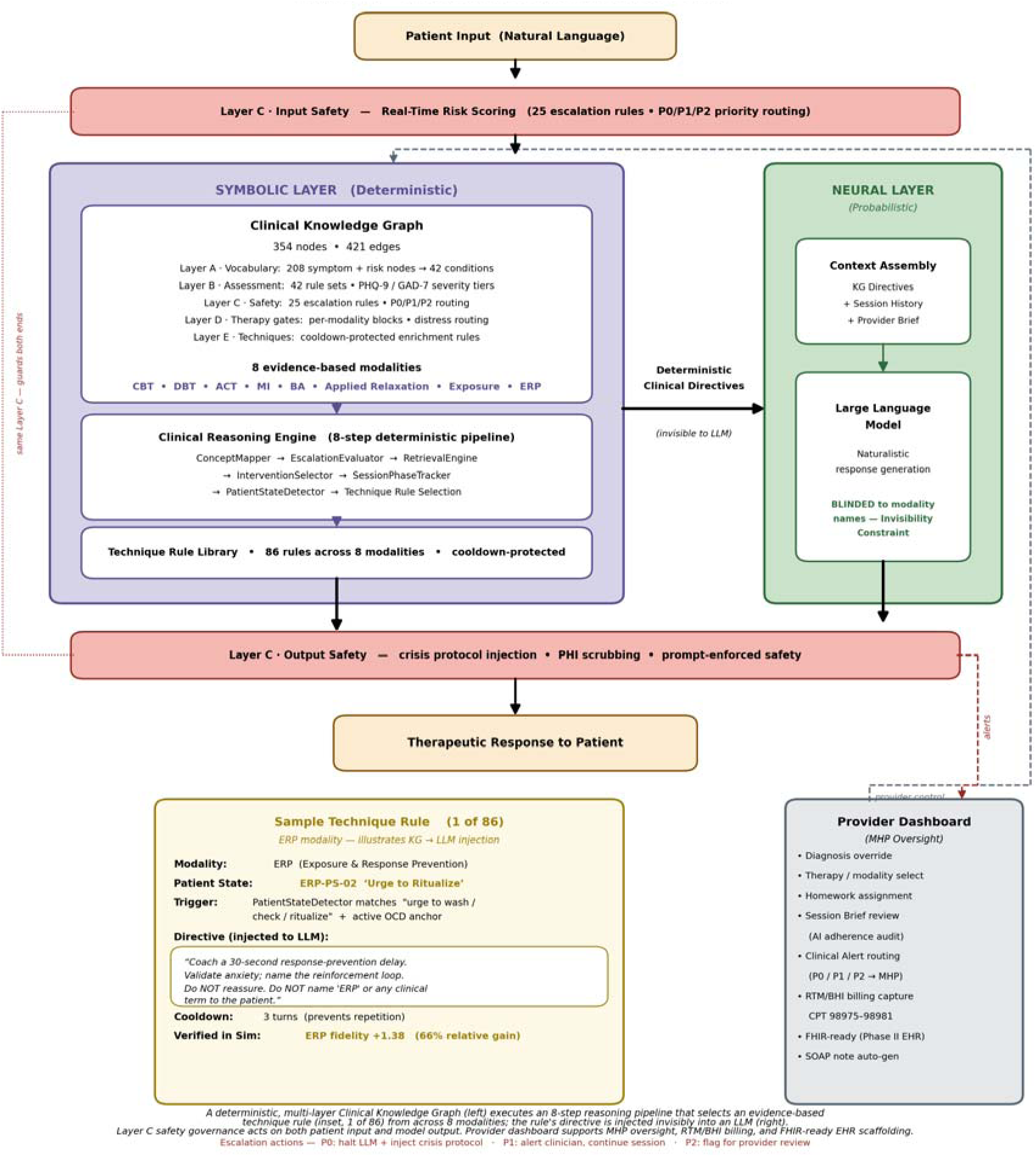
C-Mind neuro-symbolic architecture. The three-component pipeline separating clinical reasoning from language generation (Symbolic Layer → Neural Layer → Post-Response Safety Gate), detailed in §2.1; layer specifications in Supplementary Material S1.

The KG is a JSON-encoded directed graph comprising 354 nodes and 421 edges across five functional layers: vocabulary/ontology, assessment, safety, therapy gates, and techniques. It includes escalation rules capable of triggering crisis routing before therapeutic response generation and technique-application rules that direct LLM behavior at each conversational turn. Because KG-directed therapeutic content is delivered naturalistically, without naming therapy modalities or technique labels, evaluation relied on function-based scoring rather than surface keyword matching.

C-Mind operates under licensed clinician supervision. Each patient is linked to a clinician who can assign or override the working diagnosis, define treatment goals and homework, set monitoring priorities, and receive safety alerts. The provider dashboard supports validated clinical scales, including the PHQ-9, GAD-7, PCL-5, AUDIT, and C-SSRS,[26] and provides session summaries, KG activation logs, and safety audit trails.

### 2.2 Validation Design and Case Library

We evaluated simulated clinical routing, safety behavior, and governance properties of C-Mind using a pre-specified simulation-based validation framework. The evaluation addressed eight domains: clinical-domain coverage, routing correctness, safety-escalation robustness, decision repeatability, per-turn traceability, guideline-provenance auditability, treatment-goal alignment, and KG-to-LLM technique enforcement.

Two complementary simulation methods were used:

- **Vignette-based evaluation:** Complete clinical scenarios were submitted to the KG in a single pass. This method assessed routing, safety classification, and repeatability independent of progressive symptom disclosure.
- **Patient-simulation evaluation:** LLM-based patient agents disclosed information progressively across 10 conversational turns (15 in the trace-instrumented technique-enforcement analysis; §3.9), approximating deployed between-visit sessions. This method assessed turn-level governance behavior under controlled conversational conditions and was not intended to estimate real-world clinical effectiveness or patient outcomes.

The validation library included 117 standardized clinical vignettes spanning five categories (Table 1). Each vignette was anchored to a named evidence-based guideline and paired with a rubric specifying expected therapies, acceptable alternatives, contraindicated approaches, and required safety protocols. The 93 cases in the primary counseling domain—depression, anxiety, comorbid depression/anxiety, and obsessive-compulsive disorder—were additionally converted into patient-simulation agents. Diagnostic labels and therapy-modality names were removed from patient-agent profiles to reduce leakage of the expected diagnosis or treatment pathway. Case libraries, scoring criteria, analysis subsets, KG version, model versions, and system prompts were finalized before each corresponding analysis.

**Table 1.**
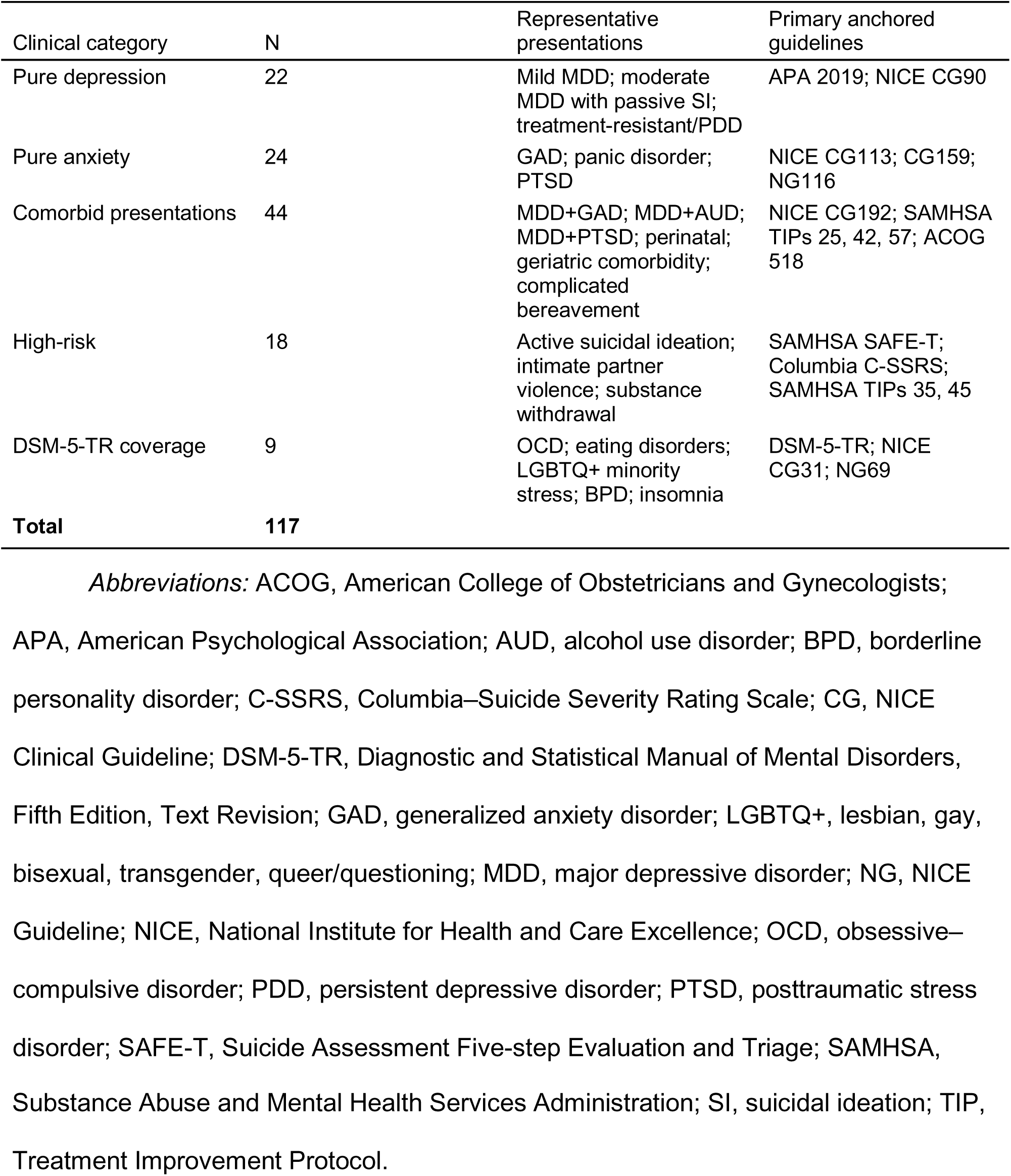
Composition of the 117-case validation library.

| Clinical category | N | Representative presentations | Primary anchored guidelines |
| --- | --- | --- | --- |
| Pure depression | 22 | Mild MDD; moderate MDD with passive SI; treatment-resistant/PDD | APA 2019; NICE CG90 |
| Pure anxiety | 24 | GAD; panic disorder; PTSD | NICE CG113; CG159; NG116 |
| Comorbid presentations | 44 | MDD+GAD; MDD+AUD; MDD+PTSD; perinatal; geriatric comorbidity; complicated bereavement | NICE CG192; SAMHSA TIPs 25, 42, 57; ACOG 518 |
| High-risk | 18 | Active suicidal ideation; intimate partner violence; substance withdrawal | SAMHSA SAFE-T; Columbia C-SSRS; SAMHSA TIPs 35, 45 |
| DSM-5-TR coverage | 9 | OCD; eating disorders; LGBTQ+ minority stress; BPD; insomnia | DSM-5-TR; NICE CG31; NG69 |
| **Total** | **117** |  |  |

*Abbreviations:* ACOG, American College of Obstetricians and Gynecologists; APA, American Psychological Association; AUD, alcohol use disorder; BPD, borderline personality disorder; C-SSRS, Columbia–Suicide Severity Rating Scale; CG, NICE Clinical Guideline; DSM-5-TR, Diagnostic and Statistical Manual of Mental Disorders, Fifth Edition, Text Revision; GAD, generalized anxiety disorder; LGBTQ+, lesbian, gay, bisexual, transgender, queer/questioning; MDD, major depressive disorder; NG, NICE Guideline; NICE, National Institute for Health and Care Excellence; OCD, obsessive–compulsive disorder; PDD, persistent depressive disorder; PTSD, posttraumatic stress disorder; SAFE-T, Suicide Assessment Five-step Evaluation and Triage; SAMHSA,
Substance Abuse and Mental Health Services Administration; SI, suicidal ideation; TIP, Treatment Improvement Protocol.

### 2.3 Structural and Adversarial Validation Without Patient Simulation

We first evaluated the KG and safety pipeline directly using single-pass vignettes, adversarial probes, and static KG audits. This analysis did not invoke patient simulation.

- **Routing validation:** The deployed C-Mind backend was invoked with cross-session memory disabled so that each vignette began from an identical clean state. Outputs were scored against the pre-specified rubric for diagnostic accuracy, therapy-class match, and safety routing. Safety routing was considered correct when the system fired a protocol at least as protective as the expected safety tier or appropriately withheld escalation when the rubric specified differentiated assessment without escalation.
- **Decision repeatability:** Repeatability was evaluated at the symbolic-reasoning level across independent runs. Three KG decision outputs were captured: disorder classification, therapy recommendation, and safety routing. LLM response generation was not invoked; therefore, repeatability results apply to clinical decision logic rather than generated counselor text. As a reproducibility control, the symptom extractor could additionally be run in consensus mode—k independent extractions per case combined by majority-vote symptom selection—a validation setting that is not enabled in deployment.
- **Guideline-provenance auditability:** A deterministic extractor assessed whether KG nodes and escalation rules contained machine-readable citation fields.
- **Crisis-detection robustness:** Adversarial single-turn probes and multi-turn conversational arcs tested high-risk disclosures, ambiguous wording, and context-dependent safety events. A probe was classified as detected if the safety pipeline emitted a CRITICAL or ELEVATED event through either KG-anchored detection pathway. The adversarial corpus (277 probes) was generated per class from C-SSRS-anchored construct definitions and clinically reviewed; probe classes, ground-truth labels, and the versioned corpus are provided in Supplementary Material S6. External validation replayed each CRADLE Bench test dialogue’s patient turns in order over a live session, scoring a detection when any safety layer fired at or before the last annotator-confirmed crisis turn. Additional scoring rules and audit specifications are provided in Supplementary Materials.

### 2.4 Patient-Simulation Governance Analyses

Five pre-specified governance analyses used patient simulation to assess turn-level behavior under controlled conversational conditions. Because these analyses required turn-by-turn instrumentation or clinician review, each used a pre-specified subset stratified by diagnosis and severity rather than the full simulation pool. High-risk cases were excluded from multi-turn therapy-delivery analyses because their expected governed behavior was deterministic crisis routing, which was evaluated through routing validation and adversarial crisis testing. Cases outside C-Mind’s depression-and-anxiety therapeutic scope were also excluded from patient-simulation governance analyses.

- **Per-turn traceability:** Simulated 10-turn sessions spanning depression, anxiety, comorbidity, and passive suicidal ideation were instrumented with a trace logger. Clinical reviewers reconstructed the reasoning pathway from patient utterance through symptom detection, disorder resolution, therapy ranking, technique selection, and safety routing using the trace as the evidence source.
- **Treatment-goal alignment:** Cold-start sessions were compared with provider-anchored sessions in which the patient record was seeded with provider-set diagnoses, treatment goals, preferred modalities, and, where applicable, safety plans. Outcomes included provider-intent transmission, diagnostic governance, safety governance, and modality alignment with the provider’s preferred modality set.
- **KG-to-LLM technique enforcement:** In a trace-instrumented run using 15-turn sessions, KG per-turn directives were joined with independently classified counselor responses. Adherence was assessed using pre-specified modality-family and clinical-function scoring instruments, including function-level fidelity scored with a CTRS-R-anchored instrument.[27]
- **KG soft-enforcement baseline:** The full C-Mind pipeline was compared with an otherwise identical KG-off condition in which the LLM operated without KG context.
- **Ungoverned comparator and regulated-conduct audit:** To approximate a consumer-facing chatbot configuration, the same patient agents interacted with the same underlying language model without the C-Mind stack: no C-Mind system prompt, patient chart, KG context, or safety infrastructure. Counselor turns across the KG-on, KG-off, and ungoverned conditions were audited for regulated-conduct behaviors, including delivering a diagnosis, providing medication advice, making autonomous treatment recommendations, recommending clinician referral, and providing crisis redirection. The primary audit was performed by an independent cross-family judge model, Claude Sonnet 4.5, using pre-specified definitions. A second-family judge model, GPT-5.4, independently evaluated a 60-turn subsample to assess inter-judge reliability. Judge instruments, agreement metrics, and transcripts are provided in Supplementary Material S5.

### 2.5 Provider–Patient Dyad Loop Verification

Separately from the simulation analyses, we conducted a team-member role-play verification of the deployed provider–patient dyad workflow using the live C-Mind backend and provider-dashboard API. One team member acted as the patient and completed scripted safety scenarios, while another acted as the provider-user and monitored and acknowledged dashboard alerts. The test verified that safety events could be detected, routed to the supervising-provider dashboard, acknowledged, and documented with patient-activity and session-time summaries. Three scenarios were tested: mid-session crisis escalation, passive suicidal ideation, and intimate-partner violence. This analysis evaluated the technical operation of the dyadic governance workflow, not provider clinical decision-making or patient clinical outcomes; results are reported in §3.8.

## 3. Results

Results are organized to mirror the pre-specified validation domains. Table 2 summarizes what each analysis tested, the data it used, and its principal result; the subsections that follow report each in turn.

**Table 2.** Evaluation overview: what each analysis tested and its principal result.

| Section | What it tests | Data / method | Principal result |
| --- | --- | --- | --- |
| §3.1 Domain coverage | KG covers the target disorders | DSM-5-TR structural audit | All named comorbidities covered |
| §3.2 Guideline routing and safety escalation | Correct routing; crisis detection and suppression | 117 vignettes (18 high-risk, 99 non-crisis) | 116/117 routed (99.1%); 18/18 crises detected, 16/18 hard-halted |
| §3.3 Adversarial robustness and external validation | Disguised or indirect risk; real crisis dialogues | 277 adversarial probes; CRADLE Bench (600 dialogues) | 96.7% sensitivity / 95.4% specificity; 98.5% detected externally |
| §3.4 Decision repeatability | Run-to-run reproducibility | 5 full-library runs | 99.1% verdict-repeatable |
| §3.5 Per-turn traceability | Reasoning reconstructable from the trace | 50 instrumented traces | 100% reconstructable |
| §3.6 Guideline-provenance auditability | Machine-readable citation coverage | 354 KG nodes and 25 escalation rules | 100% cited |
| §3.7 Provider treatment-goal governance | Provider input governs behavior | 50 provider-anchored turns | Deterministic diagnosis and safety override |
| §3.8 Provider–patient alert loop | Alerts reach and are acknowledged by the provider | Live role-play, 3 scenarios | 12 of 12 steps passed |
| §3.9 Technique enforcement and comparators | KG-directed technique is delivered | 105 turns; KG-on/off and ungoverned arms | 100% session-level delivery; governance contrast |

### 3.1 Domain Coverage

Before evaluating routing performance, we assessed whether the Knowledge Graph (KG) structurally covered the target clinical domain. Against DSM-5-TR, the KG carried dedicated diagnostic nodes for the core depressive and anxiety-spectrum disorders targeted by the platform; every named comorbidity was represented through a detection, treatment, or referral pathway; and 15 psychosocial-context conditions relevant to depression and anxiety presentations were instantiated. Of the 10 distinct comorbid disorders named in the DSM-5-TR Depressive Disorders and Anxiety Disorders chapters, seven had dedicated diagnostic nodes and three were covered through detection or differential pathways. No named comorbid condition was unhandled.

### 3.2 Guideline-Concordant Routing and Safety Escalation

Across the 117 standardized clinical vignettes, C-Mind routed 116 cases to guideline-appropriate care (99.1%; 95% CI, 95.3%–100%), exceeding the pre-specified ≥80% target. Of the 116 passing cases, 70 were scored OPTIMAL (59.8%) and 46 ACCEPTABLE (39.3%). Routing performance was consistent across clinical families, with 100% pass rates for mild depression, severe depression with passive suicidal ideation, posttraumatic stress disorder, active suicidal ideation, intimate partner violence, and substance withdrawal. The final results reported here reflect the locked system configuration used for the pre-specified evaluation.

#### High-risk (crisis) vignettes

The 18 high-risk vignettes tested whether the KG routed presentations requiring urgent attention to protective safety protocols and, where appropriate, suppressed therapeutic response generation. All 18 high-risk presentations were detected and routed to a protective response (sensitivity, 18/18; 95% CI, 81.5%–100%), and a therapy-suppressing hard halt fired in 16 of 18: 6 of 6 active suicidal ideation cases, 6 of 6 acute substance-related presentations, and 4 of 6 intimate-partner-violence presentations. The two intimate-partner-violence presentations that did not hard halt—a same-sex intimate-partner-violence pattern and elder abuse—were detected and routed to supportive safety handling at the ACCEPTABLE tier rather than emergency halt. These under-recognized patterns were flagged for clinician-in-the-loop review and targeted KG expansion.

Safety routing was scored correct in 17 of 18 high-risk cases. The sole exception—also the single non-passing case in the overall routing analysis—was a stimulant-induced-psychosis vignette in which the hard halt fired consistently, but the specific crisis-resource protocol attached to the halt varied across runs, affecting the scored verdict. In each hard-halted case, therapeutic response generation was suppressed and the supervising provider received a crisis alert within the same conversational turn; the LLM did not participate in the safety-routing decision. End-to-end traces for two illustrative cases—intimate-partner violence during pregnancy and acute alcohol withdrawal—are provided in Supplementary Material S2.

#### Non-crisis vignettes

Among the 99 non-high-risk cases, no case failed safety scoring under the conservative rubric. Protective over-escalation occurred in 11 cases (11.1%), all involving severe presentations or risk-adjacent content, including eating-disorder medical risk, borderline-personality self-harm content, minority-stress presentations with risk content, and severe comorbidity. These events were classified as conservative safety behavior under the pre-specified rubric rather than unsafe under-escalation, although such protective over-escalation may increase provider alert burden in deployed use.

### 3.3 Adversarial Robustness and External Validation

#### Adversarial robustness

Complete-information vignettes established safety behavior when risk was stated directly. A complementary adversarial stress test assessed whether the system detected indirect, disguised, or context-dependent risk language. Across 277 single-turn probes spanning nine ground-truth classes plus three multi-turn conversational arcs, whole-system sensitivity was 96.7% (95% CI, 91.9%–98.7%) and specificity was 95.4% (95% CI, 90.3%–97.9%) (Table 3; Figure 2A). Detection was complete for explicit active ideation (40/40) and for probes pairing an explicit denial with a co-present current disclosure (12/12), the minimize-then-disclose pattern by which ambivalent patients commonly surface risk. All three multi-turn arcs were handled correctly, including a five-turn benign lead-in followed by an explicit disclosure that fired on exactly the disclosure turn.

**Figure 2.**
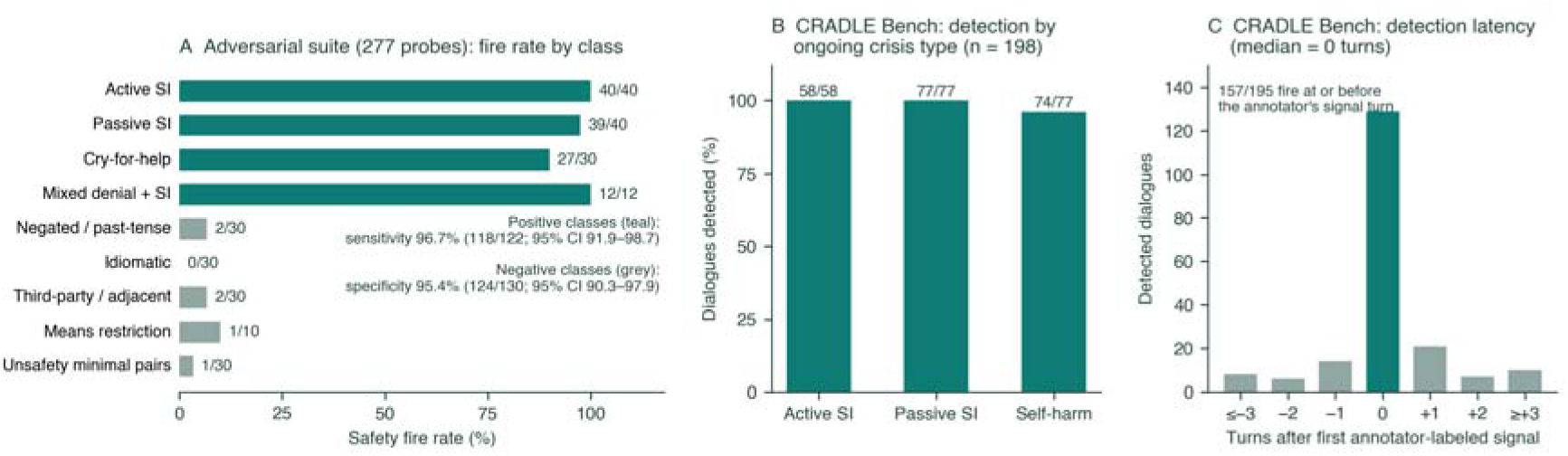
Adversarial stress testing and external validation of crisis detection. (A) Fire rate by probe class on the adversarial suite (Table 3): positive classes (teal) must fire; negative classes (grey) must stay quiet. (B) External validation on the CRADLE Bench test split: detection rate among dialogues with annotator-confirmed ongoing suicidal ideation or self-harm, by crisis type. (C) Detection latency relative to the first annotator-labeled crisis signal. Full statistics are reported in §3.3; CRADLE Bench labels follow the corpus’s Alert-Confirm protocol.[28]

**Table 3.** Adversarial suicide-risk stress test: fire rate by probe class (N = 277).

| Probe class | Construct probed | Ground truth | n | Fired | Rate |
| --- | --- | --- | --- | --- | --- |
| Active suicidal ideation | current intent, plan, or means access | Positive | 40 | 40 | 100% |
| Passive suicidal ideation | death wish without intent | Positive | 40 | 39 | 98% |
| Pre-emptive cry-for-help | fear of one's own imminent self-harm | Positive | 30 | 27 | 90% |
| Denial with co-present disclosure | benign clause + genuine current risk | Positive | 12 | 12 | 100% |
| Negated / past-tense | resolved history, hypothetical coping | Negative | 30 | 2 | 7% |
| Idiomatic / hyperbolic | figurative death language | Negative | 30 | 0 | 0% |
| Third-party / SI-adjacent | others' suicide, grief, mortality fear | Negative | 30 | 2 | 7% |
| Means restriction / coping | protective behavior disclosure | Negative | 10 | 1 | 10% |
| Unsafety minimal pairs | IPV, anger, relapse-fear phrasings | Negative | 30 | 1 | 3% |
| Gray-zone (descriptive only) | ambiguous distress, no ground truth | — | 25 | 12 | 48% |

*Table note.* Probes are single-turn utterances sent to the deployed pipeline over a live session with a neutral chart; a fire is any safety-layer activation (knowledge-graph escalation □ safety middleware). Gray-zone probes carry no ground truth and are excluded from sensitivity/specificity. Unsafety minimal pairs are scored against suicide/self-harm rules only; intimate-partner-violence or substance rules firing on those probes is correct routing. CI, Wilson score interval; SI, suicidal ideation.

#### External validation on annotated crisis dialogues

External validation used CRADLE Bench, a peer-support crisis corpus with turn-level Alert-Confirm annotations.[28] The full 600-dialogue test split was replayed in conversational order against the deployed pipeline; crisis detection was scored on the corpus’s 4,527 patient turns, with C-Mind generating the intervening counselor responses. Among 198 dialogues with annotator-confirmed ongoing suicidal ideation or self-harm, the system detected 195 (98.5%; 95% CI, 95.6%–99.5%), with median detection latency of 0 turns relative to the first annotator-labeled signal and detection at or before that turn in 157 of 195 dialogues (Figure 2B–C). Detection was complete for active (58/58) and passive (77/77) suicidal ideation; the three missed dialogues referenced self-harm solely through wound aftermath, without self-harm vocabulary in any turn. None of the 149 dialogues disclosing only past, resolved risk triggered a therapy-suppressing hard halt. Among 160 crisis-free dialogues, the system alerted the provider in 31 (specificity 80.6%; 95% CI, 73.8%–86.0%); each was a provider alert that notified the clinician without stopping the counseling session, not a therapy-suppressing hard halt. At the dialogue level, 301 dialogues drew at least one safety response (as-labeled precision, 65.8%; F1, 0.794); of the 103 apparent false positives, 38 were acknowledgments of disclosed past-tense risk and 34 were genuine crises of another type (e.g., interpersonal violence), leaving these 31 crisis-free dialogues as the operative over-notification set. This profile mirrors the conservative protective over-escalation observed in the vignette analysis (§3.2), raising provider alert burden without withholding care.

### 3.4 Decision Repeatability

Identical inputs produced highly repeatable clinical decisions across independent runs. The full 117-case library was run five times against the deployed pipeline with no code, model, or input changes between runs, and each run’s decision record was archived as a per-run artifact. The pass/fail verdict was identical across all five runs for 116 of 117 cases (99.1%). Per-run pass rates ranged from 99.1% to 100% (mean, 99.7%), with no case stably failing.

Repeatability varied by decision level. The top-ranked therapy was identical across all five runs for 98.3% of cases, with a mean pairwise Jaccard similarity of 0.958 across the full recommended-therapy set. Safety-outcome repeatability was 99.1%. In contrast, the top-ranked diagnostic label was identical across all five runs for 78.6% of cases. This lower diagnostic-label repeatability occurred primarily in near-tied comorbid presentations, where the ranker surfaced different members of the acceptable diagnosis set across runs. However, downstream clinical decisions remained stable: therapy selection, safety routing, and the scored verdict were unchanged in nearly all cases. In deployed use, provider-set diagnoses can further constrain this diagnostic-layer variation.

The single varying case identified the source of residual stochasticity. In the stimulant-induced-psychosis vignette, the hard halt fired in all five runs, indicating that the safety decision itself was stable. However, the specific crisis-resource protocol attached to the halt depended on whether the extractor surfaced the substance-use node, flipping the scored verdict in two of five runs. Thus, between-run variation originated in the stochastic symptom-extraction step rather than in the symbolic reasoning layer, and no variation crossed a safety boundary: every high-risk vignette received a crisis response in every run. A consensus-mode control (k = 3 extractions per case; §2.3) produced no additional repeatability gain, with verdict repeatability of 98.3% across five consensus runs.

### 3.5 Per-Turn Decision Traceability

Every reviewed clinical decision was reconstructable from the trace alone. All 50 trace records from the pre-specified traceability subset were captured without error. Two clinical reviewers reconstructed the complete reasoning pathway for every turn using the trace as the sole evidence source, including symptom detection, disorder resolution, therapy ranking, technique selection, and safety routing.

The depth of reconstruction is illustrated by a mild major depressive disorder case at turn 1. The patient’s opening statement, “I’ve been really tired the last few weeks. I can’t get excited about anything anymore,” produced a traceable chain from fatigue detection, to major depressive disorder resolution, to Cognitive Behavioral Therapy ranked first with Behavioral Activation second, with no safety overlay triggered. Reviewers could independently verify this chain against the KG’s guideline-linked rules.

Review of the 50 trace records identified four governance-relevant patterns. First, diagnostic resolution under progressive disclosure differed predictably from single-pass vignette results: mild major depressive disorder and generalized anxiety disorder anchored correctly from turn 1, whereas severe major depressive disorder with passive suicidal ideation required five turns to accumulate sufficient evidence. Second, social anxiety disorder showed diagnostic drift toward panic disorder when the patient framed distress around academic performance rather than explicit social-evaluation fear, identifying a symptom-alias gap not observed in single-pass evaluation. Third, Applied Relaxation was absent from generalized-anxiety-disorder therapy rankings despite being a NICE CG113 first-line recommendation, identifying a correctable routing gap. Fourth, one trace field whose label implied safety-state coverage reflected only condition-specific modifiers; full audit required cross-referencing multiple fields. These findings identified trace-schema refinements for subsequent system updates, including a dedicated escalation-state field.

### 3.6 Guideline-Provenance Auditability

All KG nodes and escalation rules contained machine-readable citation fields. Citation coverage was 100% across all 354 KG nodes and all nine node types: diagnoses (42/42), symptom and mental-state nodes (189/189), risk factors (19/19), therapy modalities (8/8), crisis-resource protocols (6/6), validated scale items (25/25), technique exercises (31/31), skills (28/28), and protective factors (6/6). At the rule layer, all 25 escalation rules also contained machine-readable citations.

The provenance fields included substantive source information. Symptom and diagnosis nodes contained DSM-5-TR criteria with page references; scale items stored PHQ-9 and GAD-7 item text with source page numbers; crisis-resource nodes stored SAMHSA SAFE-T source text; and escalation rules cited Columbia C-SSRS and PHQ-9 severity thresholds. The audit was implemented as a deterministic, re-runnable script that can be regenerated after any KG change. This audit confirms citation presence and machine readability; clinician-led review of citation correctness remains a complementary validation step.

### 3.7 Provider Treatment-Goal Governance

Provider input deterministically shaped C-Mind’s clinical behavior. In the provider-anchored comparison subset (5 cases × 10 turns = 50 provider-anchored turns), provider intent reached the LLM on all 50 turns analyzed. Provider input had measurable effects at the diagnostic, safety, and modality-selection layers.

At the diagnostic layer, provider-set diagnosis acted as an authoritative override. On the 21 of 50 turns (42%) in which the provider-set diagnosis diverged from the symptom-driven candidate diagnosis, the provider diagnosis governed the system’s framing and downstream routing. This allowed disorder-specific framing to be present from the first turn rather than requiring the system to infer the diagnosis over multiple turns of progressive disclosure.

At the safety layer, provider-supplied risk context increased vigilance. In the severe-major-depressive-disorder case with passive suicidal ideation, the escalation rule fired on 9 of 10 turns when provider clinical data were present, compared with 4 of 10 turns in the cold-start arm, where escalation depended only on what the patient disclosed in the current turn.

At the modality-selection layer, provider preference shifted therapeutic delivery. Session-level fidelity to the KG-directed modality increased from 62% in the cold-start arm to 76% in the provider-anchored arm, an absolute increase of 14 percentage points. These findings indicate that provider treatment goals were hard-enforced at the diagnostic layer, deterministically enforced at the safety layer, and soft-enforced at the modality-selection layer.

### 3.8 Provider–Patient Dyad Loop Verification

A team-member role-play integration test verified that the provider–patient governance loop functioned in the deployed system. Across three scripted risk scenarios, the system generated and surfaced alerts to the provider dashboard: two alerts during mid-session crisis escalation, four during passive suicidal ideation, and three during intimate-partner violence. Each alert was retrievable and acknowledgeable through the live API. Acknowledged alerts were removed from the active queue, and the provider-user could view real-time patient activity and session-time summaries. All 12 verification steps passed.

This verification confirms that the alert pathway operated as deployed, including risk detection, dashboard surfacing, provider-user acknowledgment, and documentation of patient activity and session time. It did not evaluate provider clinical decision-making or patient clinical outcomes, which require prospective human-subject evaluation (§4).

### 3.9 KG Technique Enforcement and Comparator Analyses

KG technique enforcement was evaluated through three complementary analyses: direct injected-versus-delivered adherence, clinical-function fidelity, and comparisons with KG-off and ungoverned LLM conditions.

#### Direct injected-versus-delivered adherence

A trace-instrumented technique-enforcement analysis included 105 turns across seven 15-turn simulated sessions. For each turn, the KG directive—including directed modality, detected patient state, and modality-specific phase—was joined with an independently classified counselor response (Supplementary Material S4). The KG-directed modality family was delivered in 100% of sessions, indicating complete session-level coverage of the intended therapeutic family.

Turn-level surface-label fidelity was lower and varied by modality. Among turns where the KG directed a specific technique, the counselor’s surface-classified modality matched the directed family on 56% of turns overall. ERP fidelity was 100%, whereas CBT fidelity was 56%. Engagement-stance responses, including Motivational Interviewing or supportive reflection, overlaid 63% of turns, consistent with the KG’s phase design, which emphasizes engagement before technique delivery within each session arc. Session-level phase-arc fidelity was 62% in the cold-start arm and 76% in the provider-anchored arm.

These findings distinguish session-level coverage from turn-level surface-label matching. The system delivered the KG-directed modality family in every session, but individual turns were often classified as engagement or supportive reflection because KG-directed therapeutic content was delivered naturalistically rather than by naming therapy techniques.

#### Function-level fidelity

Because surface modality labels may under-detect gently delivered therapeutic techniques, every technique-directed turn in the cold-start arm was also scored for clinical function using a CTRS-R-anchored instrument.[27] Across 95 scored turns, mean function fidelity was 2.50 of 5, with similar scores for CBT (2.52) and ERP (2.36). This level indicates moderate-depth delivery at the individual-turn level. The difference between surface-label fidelity and function-level fidelity suggests that some KG-directed technique content was delivered in a naturalistic form that was not always recognized by surface modality classifiers.

#### KG-on versus KG-off manipulation check

Compared with the KG-off baseline, activating the KG changed the structure of counselor responses more than their surface modality labels. Multi-paragraph responses decreased from 44% in KG-off to 9% in KG-on, a 35-percentage-point reduction. Open-question rate decreased from 36% to 17%, a 19-percentage-point reduction. KG-on turns more often ended with directive or concrete language (Supplementary Material S3).

At the modality-label level, between-arm differences were 9 percentage points or less for each modality. Differences primarily reflected redistribution among the most frequent labels: CBT (12% KG-on vs 21% KG-off), Motivational Interviewing (44% vs 50%), and supportive-labeled turns (22% vs 13%). ERP, the most structurally distinctive technique, was slightly more frequent under KG-on (8% vs 6%). Full modality-label distributions are provided in Supplementary Material S3.

#### Ungoverned LLM conversation arm

With identical patient agents and the same conversation protocol, the ungoverned LLM produced a multi-paragraph response on all 100 turns. Mean response length was 1,303 characters, compared with a mean patient utterance length of 145 characters and a governed-system mean response length of 204 characters. The ungoverned model also performed regulated clinical conduct that did not occur in either governed arm. Across the governed KG-on and KG-off arms, there were no instances of diagnosis delivery to the patient, medication advice, autonomous treatment recommendation, referral to outside clinicians, or redirection to crisis services. In contrast, the ungoverned LLM delivered a diagnosis to the patient in 4 turns across 3 cases, gave medication advice in 2 turns across 2 cases, and issued autonomous treatment recommendations in 3 turns across 2 cases.

The ungoverned arm also relied heavily on external redirection. It referred the patient to outside clinicians on 38% of turns, spanning 9 of 10 cases, and redirected the patient to crisis services on 15% of turns, spanning 6 cases. In the passive-suicidal-ideation case, the ungoverned model redirected to crisis lines on 6 of 10 turns. The ungoverned system had no provider-alert pathway, so its only available safety action was conversational redirection; the governed system, by contrast, paired in-conversation support with provider alerting for the same passive-suicidal-ideation presentation. Because the underlying language model and patient agents were held constant across arms, these differences are attributable to the C-Mind governance layer.

## 4. Discussion

This study evaluated C-Mind as a provider-supervised neuro-symbolic platform for between-visit depression and anxiety care. Its central contribution is the separation of clinical reasoning from language generation. The Clinical Knowledge Graph (KG) governs disorder resolution, therapeutic routing, contraindication handling, safety escalation, and technique selection, while the large language model (LLM) generates patient-facing responses within KG-authorized boundaries. To our knowledge, C-Mind is among the first KG–LLM hybrid architectures designed to bring this end-to-end, auditable governance model to provider-supervised between-visit mental health care.

Across the eight pre-specified governance domains (Table 2), C-Mind satisfied each criterion—routing presentations to guideline-appropriate care, detecting and suppressing high-risk cases, reproducing its decisions across independent runs, exposing every decision for per-turn reconstruction and machine-readable provenance, letting provider input govern behavior deterministically, and surfacing safety alerts to the provider dashboard for acknowledgment. Taken together rather than as isolated metrics, these findings indicate that C-Mind is not simply a conversational interface but a governed clinical AI infrastructure whose routing logic, safety decisions, guideline provenance, and provider oversight can be inspected.

Crisis detection is a particular strength—and a demanding one. Reliably identifying suicidal ideation from free text has been pursued for over a decade, yet classifier- and lexicon-based natural-language-processing methods have repeatedly traded sensitivity against false alarms and generalized poorly across corpora.[29,30] More recent fine-tuned transformer and large-language-model detectors have not resolved this: on the external benchmark used here, both general-purpose and fine-tuned language models achieve only moderate turn-level detection.[28] C-Mind, by contrast, identified suicidal ideation with near-complete sensitivity at high specificity on this independent, clinician-annotated corpus (§3.3), surpassing the strongest general-purpose and fine-tuned LLM baselines reported for it[28] and the sensitivity typical of classifier-based NLP detectors.[29] This advantage is architectural: rather than relying on a single classifier or one model judgment, C-Mind detects risk through a unified, multi-layer safety system—deterministic knowledge-graph risk-node matching and high-precision pattern rules for explicit statements, risk-signal scoring for passive or accumulating distress, and a session-level accumulator for gradual, cross-turn escalation—so that explicit, indirect, and slowly building risk are each caught by the layer best suited to it. The safety-critical hard halts are made deterministically and remain traceable to a named rule; the language model contributes only contextual disambiguation of negated or ambiguous statements, consistent with its exclusion from the safety-routing decision (§3.2). Because a missed or delayed crisis is the most consequential failure mode for a between-visit mental-health system, this combination of accuracy and accountability is clinically meaningful.

This distinction matters because clinical AI is increasingly expected to demonstrate transparency, human oversight, documentation, risk management, and independent reviewability.[20,21] In mental health care, these expectations are especially important because patient disclosures may involve suicide risk, intimate-partner violence, diagnostic ambiguity, medication questions, and fluctuating engagement between visits. Conversational fluency alone is therefore insufficient. A deployable between-visit AI system must be able to document what it detected, why it selected a pathway, how safety rules were applied, whether provider instructions were followed, and when the clinician was brought back into the loop. The three-arm comparison illustrates this point directly: when the same underlying language model operated without the C-Mind governance stack, it delivered diagnoses, provided medication advice, issued autonomous treatment recommendations, and relied on external redirection for safety. These behaviors did not occur in the governed C-Mind arms. The clinically meaningful difference was therefore not the LLM alone, but the architecture controlling how the LLM was used.

C-Mind also addresses a broader care-delivery problem. Outpatient mental health care remains largely organized around scheduled, billable encounters, while much of the clinical need in depression and anxiety occurs between visits. Patients may struggle with symptoms, safety concerns, treatment homework, coping skills, medication-related questions, or engagement long before the next appointment. Yet this between-visit work is often under-supported, inconsistently documented, and poorly aligned with reimbursement. A provider-supervised AI platform could make between-visit care more continuous, measurable, and clinically actionable. If implemented responsibly, such systems may help clinicians extend care beyond the encounter, reduce routine monitoring and documentation burden, improve continuity, and expand effective clinical capacity without replacing the treating provider.

The reimbursement pathway for this model remains an important implementation challenge. Behavioral Health Integration, Collaborative Care, and Remote Therapeutic Monitoring frameworks create potential mechanisms for reimbursing supervised monitoring, measurement-based care, provider review, and between-visit therapeutic activity.[22,23,26] However, these pathways are not universally covered or operationalized across payers. Coverage, documentation requirements, supervision rules, and claims adjudication may differ across Medicare, Medicaid, commercial insurers, and individual plans. C-Mind’s next implementation phase therefore cannot assume reimbursement as a solved problem. It should test payer-specific workflows, document which activities are reimbursable, identify denial patterns, and determine how practices can operationalize billing without increasing administrative burden. At a policy level, broader coverage and clearer reimbursement rules will be needed if between-visit care is to be valued as part of accountable mental health treatment rather than treated as uncompensated clinician labor.

Several findings also identify areas requiring refinement before clinical deployment. The system’s safety errors were asymmetric: it over-escalated on a minority of severe or risk-adjacent non-crisis presentations rather than under-escalating. This conservative posture is defensible in early-stage mental health AI, but it may increase provider alert burden and must be tuned carefully to avoid alert fatigue. In addition, two under-recognized intimate-partner-violence patterns, same-sex IPV and elder abuse, were detected and supported but were not routed to a therapy-suppressing halt. This illustrates that rule-based safety coverage is only as complete as the encoded clinical patterns and requires ongoing expansion. Finally, separating reasoning from generation constrains what the system decides, but not every word the LLM generates. Residual variation can still enter through symptom extraction and patient-facing wording. Post-response safety gates, PHI scrubbing, trace review, and provider oversight therefore remain necessary even in a KG-governed architecture.

This study has several limitations. No human patients received care through C-Mind in this evaluation. The findings are based on standardized vignettes, adversarial probes, LLM-based patient agents, and team-member role-play workflow verification. These methods cannot reproduce the full complexity of real patient disclosure, therapeutic alliance, crisis ambiguity, engagement, somatic presentation, clinician workflow, or payer documentation burden. The results therefore establish governance feasibility and internal validation, not clinical efficacy. In addition, the standardized case libraries were authored by the study team. Although cases were guideline-anchored and evaluated against pre-specified rubrics, future studies should use independently authored, held-out case libraries with rubrics frozen before evaluation. Several analyses also used focused subsets rather than the full case library, and repeatability was evaluated on a single model version. Future work should assess stability across LLM upgrades, prompt revisions, KG modifications, and deployment environments. In the external validation, C-Mind generated the counselor turns during replay, so the conversational context differed from the corpus’s original peer-listener replies; because suicidal-ideation and self-harm are scored on each patient turn, this difference does not affect the reported crisis-detection results. Finally, the provenance audit confirmed citation presence and machine readability, but not the full clinical correctness of every citation; clinician-led citation-correctness review remains necessary.

The next phase should move C-Mind from technological and functional readiness to human-centered clinical and implementation testing. A provider-supervised dyad pilot should evaluate whether the governance properties demonstrated in simulation hold in real care and whether patients and clinicians can use the system safely and meaningfully. Key outcomes should include feasibility, acceptability, patient engagement, provider usability, therapeutic alliance, symptom change, alert burden, documentation quality, safety workflow performance, and effects on provider workload. The pilot should also carry the reimbursement questions raised above into a live billing workflow, documenting which activities pay, how claims are adjudicated, and where payer policy blocks adoption.

If these next-stage studies are successful, C-Mind could support a broader reorganization of outpatient mental health care. Rather than limiting reimbursable care to scheduled encounters, provider-supervised between-visit AI could help make ongoing monitoring, skills reinforcement, safety detection, and treatment-plan support visible, reviewable, and potentially billable. This model could reduce clinician burden, expand care capacity for practices facing workforce shortages, and provide patients with more continuous support during the periods when symptoms and risks often emerge. Realizing that potential will require not only clinical validation, but also implementation science, payer engagement, and policy change that recognizes between-visit care as a core component of mental health treatment.

## 5. Conclusion

C-Mind represents a provider-supervised KG–LLM architecture for accountable between-visit mental health AI. By separating clinical reasoning from language generation, the platform makes therapeutic routing, safety escalation, guideline provenance, and provider oversight inspectable rather than leaving them embedded in unconstrained model output. The present simulation-based evaluation demonstrates technological and functional readiness for prospective human-centered testing, not clinical efficacy. The next step is to evaluate C-Mind in real provider–patient dyads while also testing the reimbursement and implementation pathways needed to make between-visit AI-supported care clinically sustainable. If validated in practice, this model could help shift outpatient mental health care from episodic visit-centered delivery toward continuous, supervised, and accountable support between visits.

## Data Availability

All data produced in the present study are available upon reasonable request to the authors

## Declarations

### Data Availability

The validation case library, simulation data, scoring scripts, and per-turn trace logs supporting the findings of this study are available from the corresponding author upon reasonable request. The external validation corpus (CRADLE Bench[28]) is publicly available on the Hugging Face Hub (dataset SungJoo/Cradle-Dialogue) under the Apache License 2.0. The deployed C-Mind system and its Clinical Knowledge Graph are proprietary to C-Mind LLC.

### Ethics

Simulation study using synthetic patient personas; no human subjects; IRB not required.

### Author Contributions (CRediT)

[Complete after author list is final — e.g., JT: conceptualization, methodology, software, validation, formal analysis, writing — original draft; clinical reviewers: validation (trace audit), writing — review & editing.]

### Declaration of Generative AI Use

Generative AI tools were used to assist with manuscript drafting and editing under full author direction; all clinical content, analyses, and claims were authored, verified, and approved by the authors, who take full responsibility for the content. [Adjust wording to the journal’s required format at submission.]

## Supplementary Materials

**Supplementary Material S1 — KG Layer Specifications.** Full specification of the five functional KG layers (vocabulary, assessment, safety, therapy gates, techniques), referenced from §2.1.

**Supplementary Material S2 — High-Risk Case Traces.** End-to-end per-turn case walkthroughs (intimate-partner violence during pregnancy; alcohol withdrawal) illustrating crisis-protocol routing and trace auditability, referenced from §3.2.

**Supplementary Material S3 — Modality-Label Distributions and Structural Markers.** Full per-modality label distributions and the structural-marker shifts (multi-paragraph rate, open-question rate, acknowledgment, directive, and concrete-cue language) for the KG-on and KG-off arms of the soft-enforcement baseline, referenced from §3.9.

**Supplementary Material S4 — KG Direction and Chat History.** Per-turn record joining the modality the Knowledge Graph directed (anchored modality, detected patient-state, modality-specific phase, and the injected clinical directive) with the chat history (patient message and counselor response) and the independently-classified counselor modality, for the trace-instrumented direct-adherence analysis (§3.9).

**Supplementary Material S5 — Ungoverned-LLM Arm and Regulated-Conduct Panel.** The ungoverned-arm conversation protocol, the regulated-conduct judge instrument with behavior definitions and the second-family reliability subsample (agreement 98.0%, κ = 0.76), and complete transcripts for all three arms (§2.4, §3.9).

**Supplementary Material S6 — Adversarial Crisis-Detection Corpus.** Corpus-generation method (C-SSRS-anchored construct definitions, per-class generation, clinical review), the ten-class probe table with per-class n, and a pointer to the versioned corpus JSON, referenced from §2.3 and §3.3.

